# Single-cell RNA Sequencing of Peripheral Blood Mononuclear Cells in Patients with Single Ventricle/Hypoplastic Left Heart Syndrome

**DOI:** 10.1101/2024.08.20.24312290

**Authors:** Hui-Qi Qu, Kayleigh Ostberg, Diana J Slater, Fengxiang Wang, James Snyder, Cuiping Hou, John J Connolly, Michael March, Joseph T Glessner, Charlly Kao, Hakon Hakonarson

## Abstract

**Background:** Single ventricle and hypoplastic left heart syndrome (SV/HLHS) patients require lifelong medical monitoring and management to address potential complications and optimize their health. The consequence of SV/HLHS had detrimental effects on multiple organ systems, including on peripheral blood mononuclear cells (PBMCs) and can weaken the immune system, exacerbating the risk of infection and various cardiovascular complications.

**Methods:** Using single-cell RNA sequencing (scRNA-seq), we studied PBMCs from 33 pediatric patients (10 females and 23 males) with SV/HLHS. By a pair-wide study design, the SV/HLHS patients were compared to 33 controls without heart diseases.

**Results:** Four cell types account for the top 62% cumulative importance of disease effects on gene expression in different cell types, i.e., [T cells, CD4+, Th1/17], [T cells, CD4+, TFH], [NK cells], and [T cells, CD4+, Th2]. Significant sex differences were observed in [T cells, CD4+, TFH], with less prominent effects in female patients. A total of 6659 genes in different cell types were significantly differentially expressed (DE). Hierarchical clustering by WGCNA analysis of the DE genes revealed that DE genes in NK cells are most closely related to those in SV/HLHS. A total of 822 genes showed cell specific DE with opposite directions in different cell types, highlighting overrepresented MYC and IFN-γ activity in T cell and NK cell populations, as well as underrepresentation in monocytes and Treg cells.

**Conclusion:** This study elucidates the complex transcriptome landscape in PBMCs in patients with SV/HLHS, emphasizing the differential impacts on various cell types. New insights are gained into the precise modulation of MYC and IFN-γ activity in SV/HLHS, which may help balance immune responses and reduce harmful inflammation, and promote effective tissue repair and infection control.

## 1. Introduction

Single ventricle and hypoplastic left heart syndrome (SV/HLHS) are complex congenital heart defects, necessitating restructuring of circulation to optimize blood flow and oxygenation^1^. Affected patients require lifelong medical monitoring and management to address potential complications and optimize their health. Peripheral blood mononuclear cells (PBMCs) have been implicated in the pathophysiology of various cardiovascular complications, including heart failure, arrhythmias, and vascular dysfunction. In particular, infection is a significant complication for individuals with SV/HLHS. The frequent use of central lines, catheters, and other medical devices in these patients serves as potential sources of infection^2^. In addition to bloodstream infections, patients are also at an increased risk for respiratory infections due to impaired immune function and potential respiratory complications from heart failure^3^. The chronic impact of SV/HLHS on PBMCs can weaken the immune system, exacerbating the risk of infection and various cardiovascular complications. Immunosuppression heightens susceptibility to both bloodstream and respiratory infections, further complicating clinical management. The altered immune profile contributes to cardiovascular issues such as heart failure, arrhythmias, and vascular dysfunction. Understanding the molecular and cellular mechanisms underlying these complications is crucial. Key research questions include identifying the specific gene expression changes in PBMCs of patients with SV/HLHS compared to healthy controls and elucidating the underlying molecular pathways in PBMCs that contribute to the altered immune profile in SV/HLHS patients. Transcriptome profiling of PBMCs can offer valuable insights into the pathogenesis of SV/HLHS-related complications.

Using single-cell RNA sequencing (scRNA-seq), we examined the impact of SV/HLHS on PBMCs, uncovering sex differences across various cell types. By focusing on differentially expressed (DE) genes in SV/HLHS, we identified cell type-specific expression of a number of genes related to SV/HLHS. Additionally, we highlighted co-expressed gene sets related to SV/HLHS, providing new insights into the molecular underpinnings of the disease.

## 2. Methods

### 2.1 Subjects

This study included 33 pediatric patients (10 females and 23 males) with SV/HLHS. Among these patients, 20 had European ancestry, 9 had African ancestry, 1 had Asian ancestry, and 3 had other ancestries. The ages ranged from 0 to 21 years, with a median age of 6 years. Fifteen patients were diagnosed with SV, 24 were diagnosed with HLHS, and 18 underwent the Fontan procedure. Using a pair-wise study design, these patients were compared to 10 female and 23 male de-identified children without heart disease. All experimental protocols were approved by the Institutional Review Board (IRB) of the Children’s Hospital of Philadelphia (CHOP) with the IRB number: IRB 16-013278. Informed consent was obtained from all subjects. If subjects are under 18, consent was also obtained from a parent and/or legal guardian with assent from the child if 7 years or older.

### 2.2 PBMC scRNA-seq

Blood samples were collected from each participant in EDTA-coated tubes and immediately processed at the Center for Applied Genomics (CAG) at CHOP. PBMCs were isolated using Ficoll density gradient centrifugation. scRNAseq was conducted with the 10X Chromium Single Cell Gene Expression assay (10x Genomics, Single Cell 3’ v3). Sequencing was carried out on the Illumina HiSeq2500 platform using SBS v4 chemistry. The resulting data from the Chromium single-cell RNA sequencing were processed and analyzed using Cell Ranger 7.1.0 software (10x Genomics), with reads aligned to the GRCh38 reference genome.

Each pair of subjects was considered an independent experiment. The scRNA-seq data from the 33 independent pairs of children were analyzed using the Seurat R package^4 5^ for pair-wise comparison, employing SCTransform for normalization and scaling. To enhance comparability among samples, Harmony ^6^ was used to align datasets post-principal component analysis (PCA). Uniform manifold approximation and projection (UMAP) (Becht et al. 2019) was applied to group cells into clusters. Analysis included 15 cell types, identified using singleR and the celldex::DatabaseImmuneCellExpression Data() function^7^ (Supplementary Figure 1).

### 2.3 Assessing cell types affected by SV/HLHS using machine learning

To assess the impact of SV/HLHS on different cell types, data were structured into a matrix where rows represented individual genes and subject pairs, and columns denoted different cell types. Each cell in the matrix contained the log2 fold change (log2FC) in gene expression for each cell type between each case/control pair. Missing values, due to insufficient cell numbers for a specific gene in a cell type, were imputed using the mean log2FC from other experiments. If a gene was not expressed in a particular cell type, it was assigned a value of 0.

A RandomForestRegressor from scikit-learn^8^ was selected for its ability to manage high-dimensional data and generate feature importance metrics. The dataset was split into training and testing sets with the 0.8-0.2 train-test split. The number of trees (n_estimators) was set to 100, max_depth to none, min_samples_split to 2, and random_state to 42. The model’s robustness was assessed using 5-fold cross-validation (CV) to prevent overfitting, with the mean squared error (MSE) calculated for each fold to evaluate performance. Post-training, feature importance values were used for indicating the relative contribution of each cell type to the model’s predictions. Cell types were then ranked according to their importance scores. To estimate the statistical significance of feature importance values, the permutation_importance function from sklearn.inspection was employed. This function shuffled feature values to perform 1000 permutations for each feature. The p-value for each feature was computed as the proportion of permuted importance values that were equal to or greater than the original importance values.

### 2.4 Differential expression (DE) analysis

Based on the results of the Seurat R package^4 5^, a gene with DE was defined as FDR<0.05 in at least 2 pairs of samples, and with log2FCs in the same direction within the same cell type. Distribution of DE genes by cell type is visualized with the Venn diagram and the UpSetR package^9^.

### 2.5 Weighted Gene Co-expression Network Analysis (WGCNA) of DE genes

WGCNA analysis was done in 6319 DE genes with expression detected in each of the 66 PBMC samples. The average expression level per cell type for each gene in each sample was calculated using log-normalized values with the Seurat R package^4 5^. The WGCNA analysis was performed using the WGCNA R package^10 11^. Hallmark gene set overrepresentation analysis (ORA) were done using the clusterProfiler R package^12^ and the msigdbr R package^13^.

## 3. Results

### 3.1 Cell types affected by SV/HLHS

The CV MSE scores, the mean MSEs, and the standard deviations are shown in Table 1. The test MSEs are close to the mean CV MSEs in all three groups, indicating consistent model performance. The 15 cell types along with their corresponding feature importance values are shown in Table 2. Among the 15 cell types, two cell types, i.e. [T cells, CD8+, naive, stimulated] and [T cells, CD4+, naive, stimulated], were informative in less than 50% of the subject pairs. All the other 13 cell types were informative in at least 27 (82%) out of the 33 pairs of subjects.

**Table 1.** Different parameters and train-test split ratios tested for the RandomForestRegressor.

| **Subjects** | **Cross-Validation MSE Scores** | **Mean CV MSE** | **Standard Deviation of CV MSE** | **Test MSE** |
| --- | --- | --- | --- | --- |
| All | [4.62e-03, 5.20e-03, 4.88e-03, 5.13e-03, 5.23e-03] | 5.01e-03 | 2.31e-04 | 4.81e-03 |
| Females | [6.00e-03, 5.75e-03, 6.37e-03, 6.24e-03, 5.32e-03] | 5.94e-03 | 3.72e-04 | 5.76e-03 |
| Males | [5.54e-03, 5.45e-03, 5.73e-03, 5.56e-03, 5.68e-03] | 5.59e-03 | 1.00e-04 | 5.38e-03 |

Four cell types account for the top 62% cumulative importance in disease effects on gene expression in different cell types, i.e., [T cells, CD4+, Th1/17], [T cells, CD4+, TFH], [NK cells], and [T cells, CD4+, Th2]. All four cell types have permutation P-values<0.001 in all subject pairs. Significant sex differences were observed in [T cells, CD4+, TFH], which was not affected in female patients (Table 2, Figure 1).

**Table 2.** Cell types with corresponding feature importance values.

| **Cell type** | **Importance**<br>**(all)** | **Permutation**<br>**P-value\* (all)** | **Importance**<br>**(female)** | **Permutation**<br>**P-value\***<br>**(female)** | **Importance**<br>**(male)** | **Permutation**<br>**P-value\***<br>**(male)** |
| --- | --- | --- | --- | --- | --- | --- |
| T cells, CD4+, Th1/17 | 0.2857 | 0.000 | 0.2874 | 0.000 | 0.2866 | 0.000 |
| T cells, CD4+, TFH | 0.1330 | 0.000 | 0.0491 | 0.939 | 0.1325 | 0.000 |
| NK cells | 0.1126 | 0.000 | 0.1269 | 0.000 | 0.0909 | 0.000 |
| T cells, CD4+, Th2 | 0.0860 | 0.000 | 0.0488 | 0.000 | 0.0820 | 0.000 |
| Monocytes, CD14+ | 0.0639 | 1.000 | 0.0633 | 1.000 | 0.0634 | 1.000 |
| T cells, CD4+, naïve Treg | 0.0528 | 0.000 | 0.0332 | 0.000 | 0.0832 | 0.000 |
| T cells, CD4+, Th17 | 0.0500 | 0.000 | 0.0876 | 0.000 | 0.0410 | 0.000 |
| Monocytes, CD16+ | 0.0494 | 1.000 | 0.0501 | 1.000 | 0.0484 | 1.000 |
| T cells, CD8+, naïve | 0.0320 | 1.000 | 0.0274 | 0.000 | 0.0316 | 0.984 |
| T cells, CD4+, naïve | 0.0294 | 0.000 | 0.1192 | 0.000 | 0.0262 | 0.000 |
| B cells, naïve | 0.0285 | 0.000 | 0.0328 | 0.000 | 0.0253 | 1.000 |
| T cells, CD4+, memory Treg | 0.0259 | 0.000 | 0.0229 | 0.000 | 0.0349 | 0.000 |
| T cells, CD8+, naïve, stimulated<sup>#</sup> | 0.0190 | 1.000 | 0.0194 | 1.000 | 0.0182 | 1.000 |
| T cells, CD4+, Th1 | 0.0189 | 1.000 | 0.0223 | 1.000 | 0.0206 | 1.000 |
| T cells, CD4+, naïve, stimulated<sup>#</sup> | 0.0131 | 0.000 | 0.0094 | 0.000 | 0.0151 | 0.000 |

\*Based on 1000 permutations. <sup>#</sup>[T cells, CD8+, naïve, stimulated] and [T cells, CD4+, naïve, stimulated] were informative in less than 50% of the subject pairs.

**Fig. 1.**
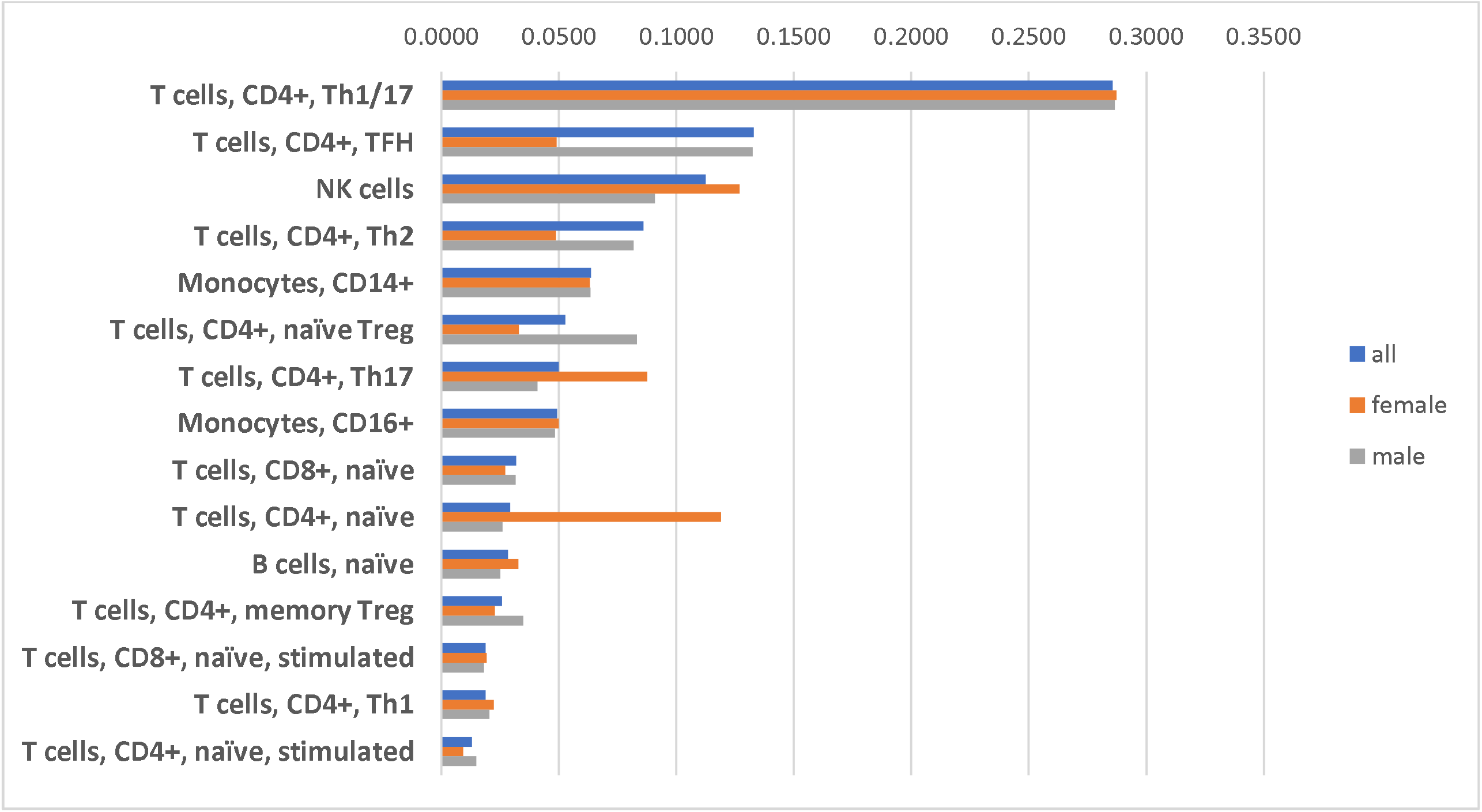
Cell types affected in SV/HLHS. X-axis represents feature importance in RandomForestRegressor.

### 3.2 DE genes

A total of 6659 genes in different cell types showed significant DE in the same direction in at least two independent pairs of samples (Supplementary Table 1). Gene sets that play critical roles in energy production, immune regulation, cell survival and proliferation, and maintenance of cellular integrity are significantly overrepresented (Table 3, Supplementary Figure 2). The distribution of the DE genes by cell type is shown in Figure 2. Among these genes, 822 genes showed opposite directions in different cell types (Supplementary Table 2).

**Table 3.** Hallmark gene sets overrepresented in differentially expressed genes in SV/HLHS.

| **HALLMARK** | **GeneRatio** | **BgRatio** | **pvalue** | **p.adjust** | **qvalue** |
| --- | --- | --- | --- | --- | --- |
| MYC_TARGETS_V1 | 164/2073 | 200/4383 | 3.64E-25 | 9.10E-24 | 5.75E-24 |
| OXIDATIVE_PHOSPHORYLATION | 164/2073 | 200/4383 | 3.64E-25 | 9.10E-24 | 5.75E-24 |
| INTERFERON_GAMMA_RESPONSE | 144/2073 | 200/4383 | 3.74E-13 | 6.23E-12 | 3.93E-12 |
| TNFA_SIGNALING_VIA_NFKB | 137/2073 | 200/4383 | 4.74E-10 | 5.92E-09 | 3.74E-09 |
| PROTEIN_SECRETION | 74/2073 | 96/4383 | 1.62E-09 | 1.62E-08 | 1.02E-08 |
| MTORC1_SIGNALING | 132/2073 | 200/4383 | 3.83E-08 | 3.19E-07 | 2.02E-07 |
| HEME_METABOLISM | 130/2073 | 200/4383 | 1.89E-07 | 1.35E-06 | 8.54E-07 |
| INTERFERON_ALPHA_RESPONSE | 67/2073 | 97/4383 | 9.69E-06 | 6.06E-05 | 3.82E-05 |
| ALLOGRAFT_REJECTION | 123/2073 | 200/4383 | 2.55E-05 | 1.41E-04 | 8.93E-05 |
| TGF_BETA_SIGNALING | 40/2073 | 54/4383 | 5.25E-05 | 2.63E-04 | 1.66E-04 |
| IL2_STAT5_SIGNALING | 120/2073 | 199/4383 | 1.12E-04 | 5.07E-04 | 3.21E-04 |
| UNFOLDED_PROTEIN_RESPONSE | 70/2073 | 113/4383 | 1.07E-03 | 4.47E-03 | 2.82E-03 |
| REACTIVE_OXYGEN_SPECIES_PATHWAY | 34/2073 | 49/4383 | 1.39E-03 | 5.35E-03 | 3.38E-03 |
| APOPTOSIS | 94/2073 | 161/4383 | 2.63E-03 | 9.39E-03 | 5.93E-03 |
| ANDROGEN_RESPONSE | 61/2073 | 100/4383 | 3.71E-03 | 1.24E-02 | 7.80E-03 |
| DNA_REPAIR | 87/2073 | 150/4383 | 4.82E-03 | 1.51E-02 | 9.52E-03 |
| COMPLEMENT | 112/2073 | 200/4383 | 7.15E-03 | 2.10E-02 | 1.33E-02 |
| ADIPOGENESIS | 110/2073 | 200/4383 | 1.54E-02 | 4.21E-02 | 2.66E-02 |
| PI3K_AKT_MTOR_SIGNALING | 61/2073 | 105/4383 | 1.60E-02 | 4.21E-02 | 2.66E-02 |
| MITOTIC_SPINDLE | 109/2073 | 199/4383 | 1.84E-02 | 4.59E-02 | 2.90E-02 |

**Fig. 2.**
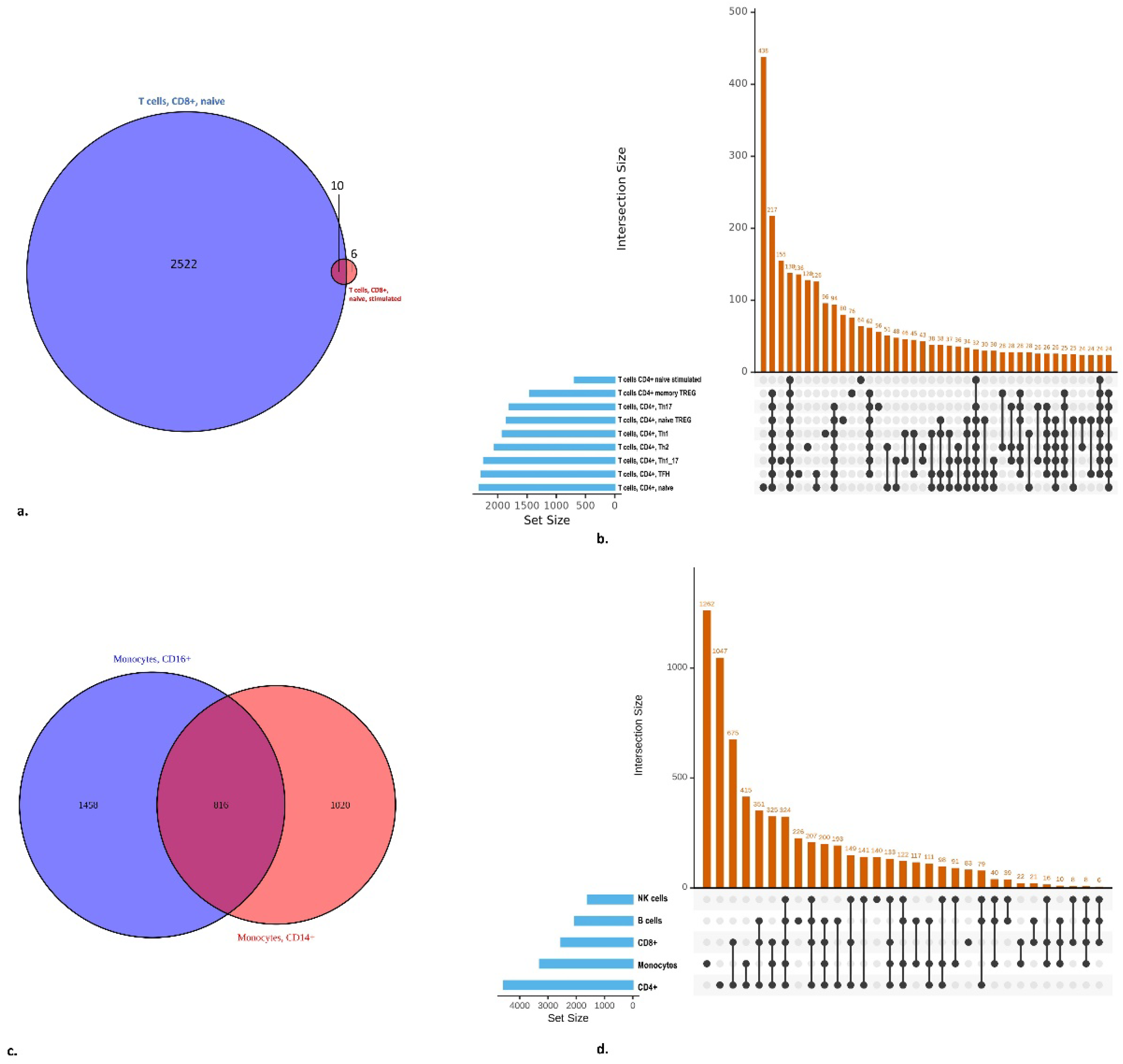
Distribution of DE genes by cell type

### 3.3 WGCNA of DE genes

Among the 6659 DE genes, 6319 genes have expression detected in each of the 66 PBMC samples. Among the 15 cell types, 9 cell types are informative in all the 66 PBMC samples, including [Monocytes, CD14+], [NK cells], [T cells, CD4+, memory Treg], [T cells, CD4+, naïve], [T cells, CD4+, TFH], [T cells, CD4+, Th1], [T cells, CD4+, Th1/17], [T cells, CD4+, Th17], and [T cells, CD4+, Th2]. The genes were grouped into different modules based on the similarity of their expression patterns across samples (Supplementary Table 3). The Hallmark gene sets highlighted in each module are shown in Supplementary Table 4. The correlations of module eigengenes with cell types, sex, and disease status are shown in Fig.3. Among these modules, the turquoise and pink modules are significantly overexpressed in SV/HLHS. The turquoise module is significantly overexpressed in [NK cells] (Fig.3). Two Hallmark gene sets, MITOTIC_SPINDLE and G2M_CHECKPOINT, are highlighted in the turquoise module (Table 4). Genes in the pink module exhibited variable expression patterns in relation to cell types and disease correlation. Specifically, these genes showed higher expression in [Monocytes, CD14+], but their expression in [Monocytes, CD14+] was downregulated in SV/HLHS. Additionally, their expression in T cells were overexpressed in SV/HLHS. No Hallmark genes were significantly overrepresented within this gene set.

**Fig. 3.**
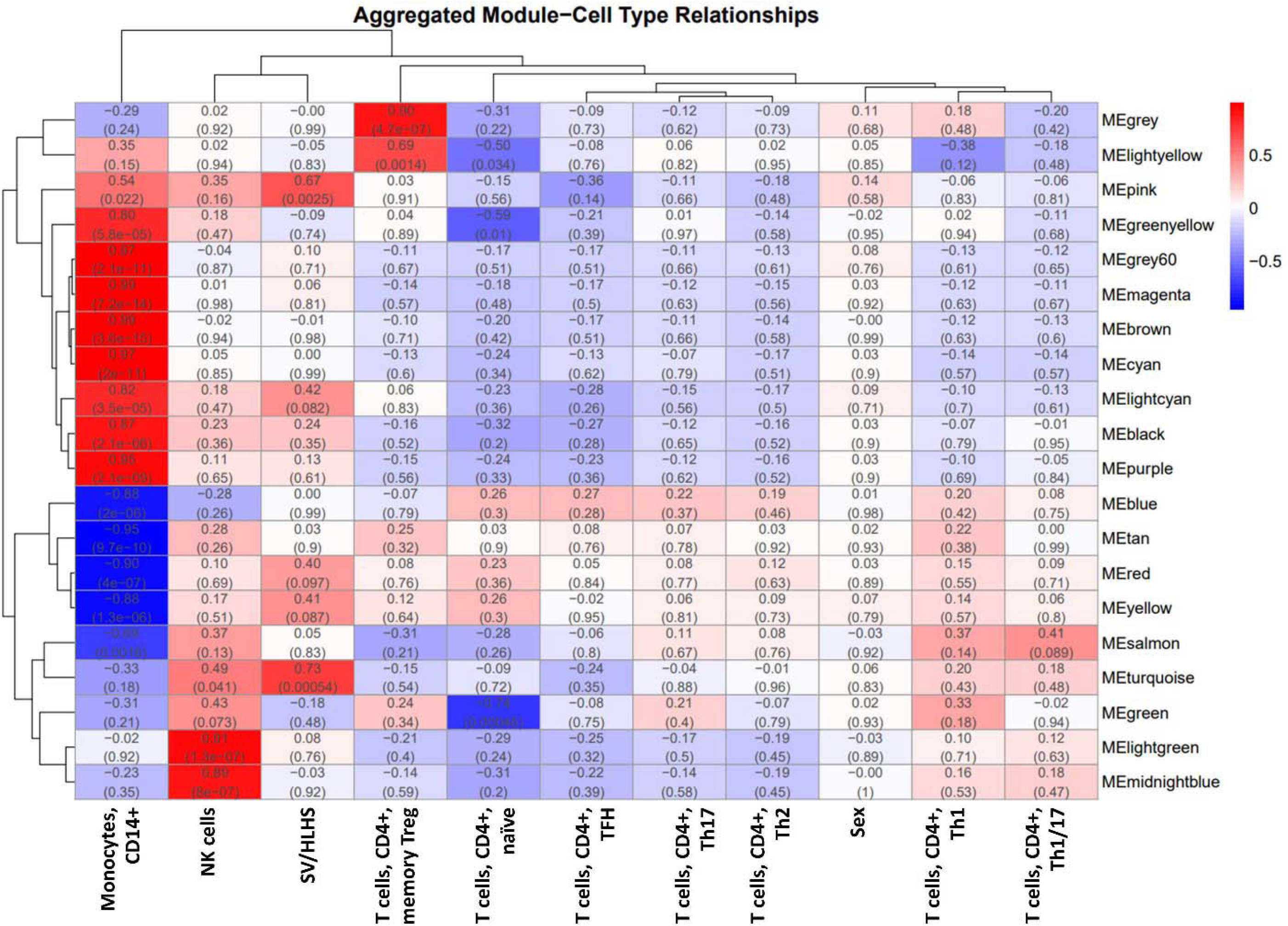
WGCNA module relationships

**Table 4.**
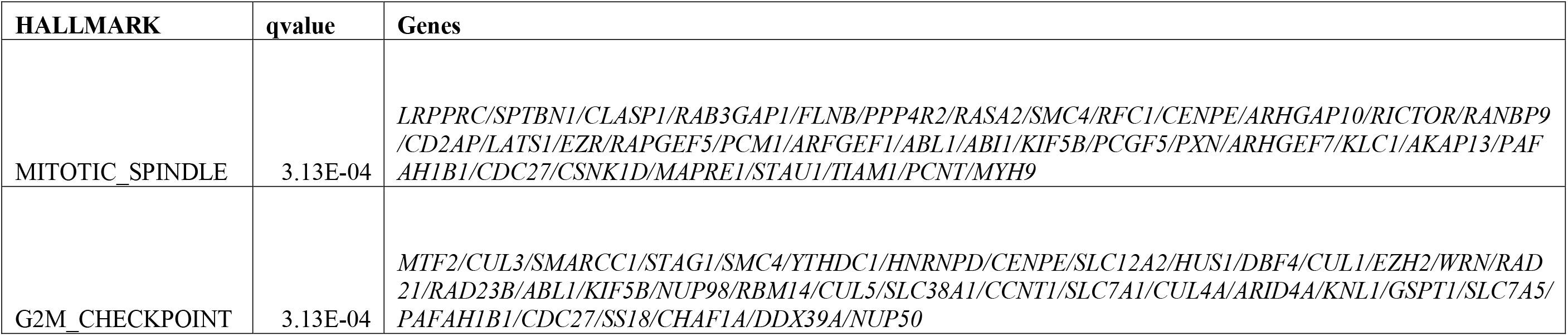
Hallmark gene sets overrepresented in the turquoise module correlated with SV/HLHS and NK cells.

## 4. Discussion

### 4.1 Immune cells affected by SV/HLHS

This study showed four cell types of T cells to be most affected in SV/HLHS, i.e., [T cells, CD4+, Th1/17], [T cells, CD4+, TFH], [NK cells], and [T cells, CD4+, Th2]. Th1/17 cells producing both IFN-γ (Th1) and IL-17 (Th17). These cell types and cytokines may help combat infections, but may also contribute to immune dysregulation involved in myocardial damage, fibrosis, and vascular dysfunction^14^. In these cells, the Interferon-Induced Proteins with Tetratricopeptide Repeats (IFIT) family of proteins, including *IFIT1, IFIT1B, IFIT2*, and *IFIT3*, are significantly overexpressed in SV/HLHS, indicating an upregulated response to interferon signaling^15^. TFH cells are critical for the formation of germinal centers and the production of high-affinity antibodies^16^. The activation of TFH cells in SV/HLHS may be due to recurrent infections and chronic inflammation. Their prolonged activation can exacerbate cardiovascular complications, increasing the risk of autoimmunity. Interestingly, our study showed that TFH cells in females were less affected, which may explain their better clinical outcomes. Th2 cells are involved in the immune responses, including regulation of allergic responses^17^. Chronic activation of these cell types can contribute to an inflammatory environment and exacerbate immune dysregulation.

### 4.2 NK cells and related DE genes

This study identified 6659 DE genes, which are overrepresented with critical roles in energy production, immune regulation, cell survival and proliferation, and maintenance of cellular integrity. By WGCNA analysis of the DE genes, hierarchical clustering revealed that genes expressed in NK cells are most closely related to SV/HLHS (Fig.3). NK cells are a vital component of the innate immune system, targeting and killing virally infected cells^18^. Genes in the lightgreen (HALLMARK_TNFA_SIGNALING_VIA_NFKB overrepresented in this module, with higher expression in NK cells, upregulated in SV/HLHS), midnightblue (HALLMARK_ALLOGRAFT_REJECTION overrepresented in this module, with higher expression in NK cells, downregulated in SV/HLHS), and turquoise (HALLMARK_MITOTIC_SPINDLE and HALLMARK_G2M_CHECKPOINT overrepresented in this module, with higher expression in NK cells, upregulated in SV/HLHS) modules were significantly correlated with NK cells (Supplementary Table 4). Upregulation of genes involved in the TNF-alpha signaling pathway via the NF-kB pathway regulates cell proliferation, differentiation, and survival, and leads to the activation of inflammatory responses^19^. The aggregate effects of downregulated expression of the gene set HALLMARK_ALLOGRAFT_REJECTION in NK cells may result in reduced cytotoxic activity and impaired ability to target and eliminate infected cells, weakening overall immune surveillance and response^13^. The overexpression of the genes of HALLMARK_MITOTIC_SPINDLE and HALLMARK_G2M_CHECKPOINT in NK cells can increase proliferation and cell cycle activity^20^.

### 4.3 Cell type specific DE genes

Cell-type specific DEs are commonly identified in this study, underscoring cellular heterogeneity consequent to SV/HLHS (Fig.2). In particular, 822 (12%) genes showed cell-type specific DE with opposite directions in different cell types (Supplementary Table 2). Among these, a number of genes are related to MYC targets, variant 1 and interferon gamma response.

The HALLMARK_MYC_TARGETS_V1 genes are direct targets of the MYC transcription factor, which is a critical regulator of cell growth, proliferation, and metabolism^21^. In SV/HLHS, these genes tend to be upregulated in [T cells, CD4+, naive], [T cells, CD4+, TFH], [T cells, CD4+, Th1], and [T cells, CD8+, naive]; and downregulated in [Monocytes, CD14+], [Monocytes, CD16+], [T cells, CD4+, memory TREG], [T cells, CD4+, naive, stimulated], [T cells, CD4+, Th17], and [T cells, CD4+, Th2]. The differential regulation of MYC targets underscores the functional specialization between T cells and monocytes. The upregulation in naive CD4+ T cells, TFH cells, Th1 cells, and naive CD8+ T cells, particularly upon activation, require robust proliferative capacity, which is supported by MYC target upregulation^22^. In contrast, monocytes, which are more involved in immediate and direct immune responses, do not rely heavily on such proliferation mechanisms^23^. The downregulation in memory Treg cells, which are crucial for maintaining immune tolerance and preventing autoimmunity^26^, may lead to a reduced number for these cells to control excessive immune responses, possibly leading to autoimmunity or prolonged inflammation.

The HALLMARK_INTERFERON_GAMMA_RESPONSE genes are upregulated in response to interferon-gamma (IFN-γ), a cytokine crucial for innate and adaptive immunity^24^. In SV/HLHS, these genes tend to be upregulated in [T cells, CD4+, naive], [T cells, CD4+, naive, stimulated], [T cells, CD4+, TFH], [T cells, CD4+, Th17], [T cells, CD4+, Th1], and [T cells, CD8+, naive]; and downregulated in [Monocytes, CD14+] and [Monocytes, CD16+]. The upregulation of IFN-γ response genes in T cells suggests a robust activation state and readiness for immune response in these cells, indicating a potential for strong pro-inflammatory signaling. Conversely, monocytes play a crucial role in inflammation and tissue repair^23^. Monocytes differentiate into macrophages and dendritic cells, which help clear debris, secrete growth factors, and regulate inflammation, thus promoting healing and regeneration^25^. The reduced IFN-γ response in monocytes might imply a diminished capacity to manage inflammation and repair tissue^23^.

Precisely modulating MYC and IFN-γ activity in SV/HLHS, based on the insights gained from our study, may balance immune responses, reduce harmful inflammation, and promote effective tissue repair and infection control. More specifically, at the cellular level, this precise modulation could involve targeting overactive T cell populations while bolstering the proliferation and function of monocytes, balancing effective infection control with minimizing harmful inflammation.

## Conclusion

This study elucidates the complex transcriptome landscape in PBMCs in SV/HLHS, emphasizing the differential impacts on various cell types. The significant correlation between gene expression in NK cells and SV/HLHS highlights a cellular emphasis. A number of gene sets in NK cells offer insights into the underlying mechanisms and provide targets for intervention in SV/HLHS. Additionally, specific genes that exhibit differential expression in opposite directions across various cell types present unique targets and opportunities to fine-tune immunity in SV/HLHS. Future research should explore therapeutic interventions aimed at modulating immune responses by targeting the critical gene sets identified in this study.

## Data Availability

All data produced in the present study are available upon reasonable request to the corresponding author.

## Ethics Approval and Consent to Participate

All experimental protocols were approved by the Institutional Review Board (IRB) of the Children’s Hospital of Philadelphia (CHOP) with the IRB number: IRB 16-013278. Informed consent was obtained from all subjects. If subjects are under 18, consent was also obtained from a parent and/or legal guardian with assent from the child if 7 years or older.

## Funding

The study was supported by the Institutional Development Funds from the Children’s Hospital of Philadelphia to the Center for Applied Genomics, and The Children’s Hospital of Philadelphia Endowed Chair in Genomic Research to HH.

**Supplementary Table 1 Average log2FC of the DE genes by the 15 cell types**

*Positive value means higher expression in cases.

**Supplementary Table 2 DE genes with opposite directions in different cell types**

*Positive value means higher expression in cases.

**Supplementary Table 3 The DE genes grouped into different modules**

**Supplementary Table 4 Significant Hallmark gene sets overrepresented in each module**

**Supplementary Figure 1.**
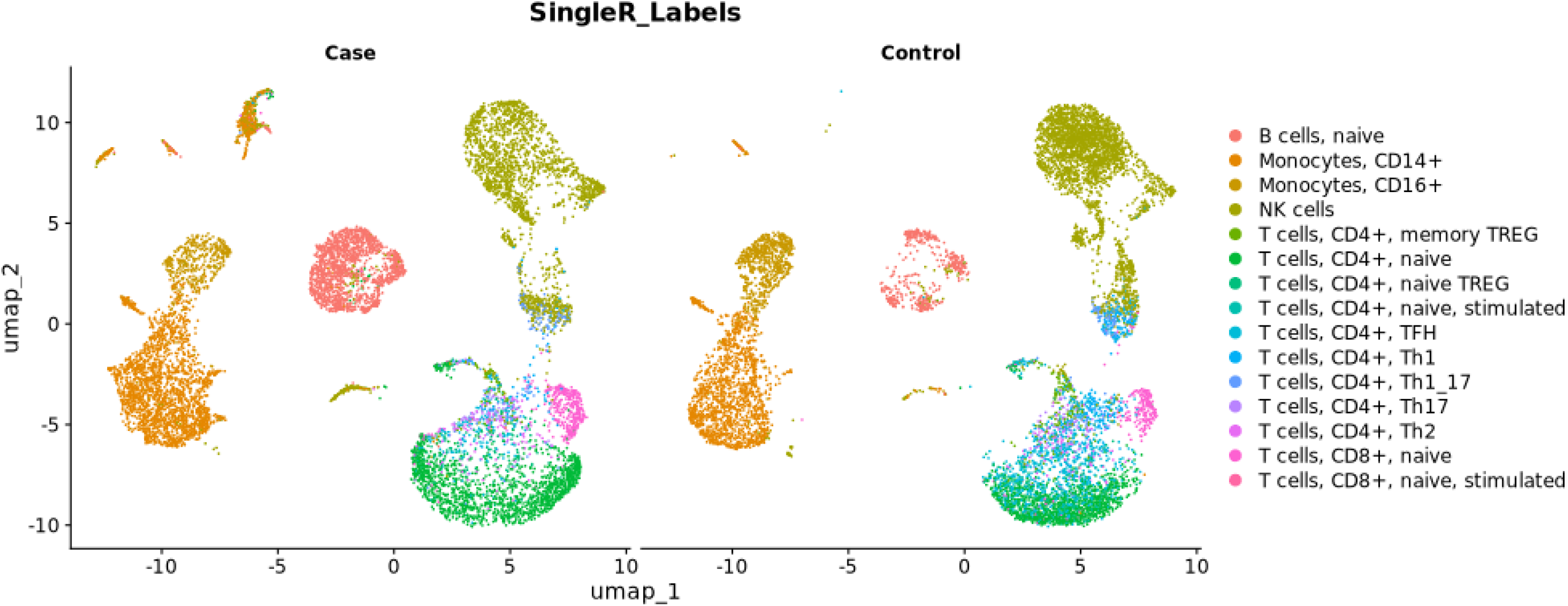
Cell types of PBMCs in a case and control pair.

**Supplementary Figure 2.**
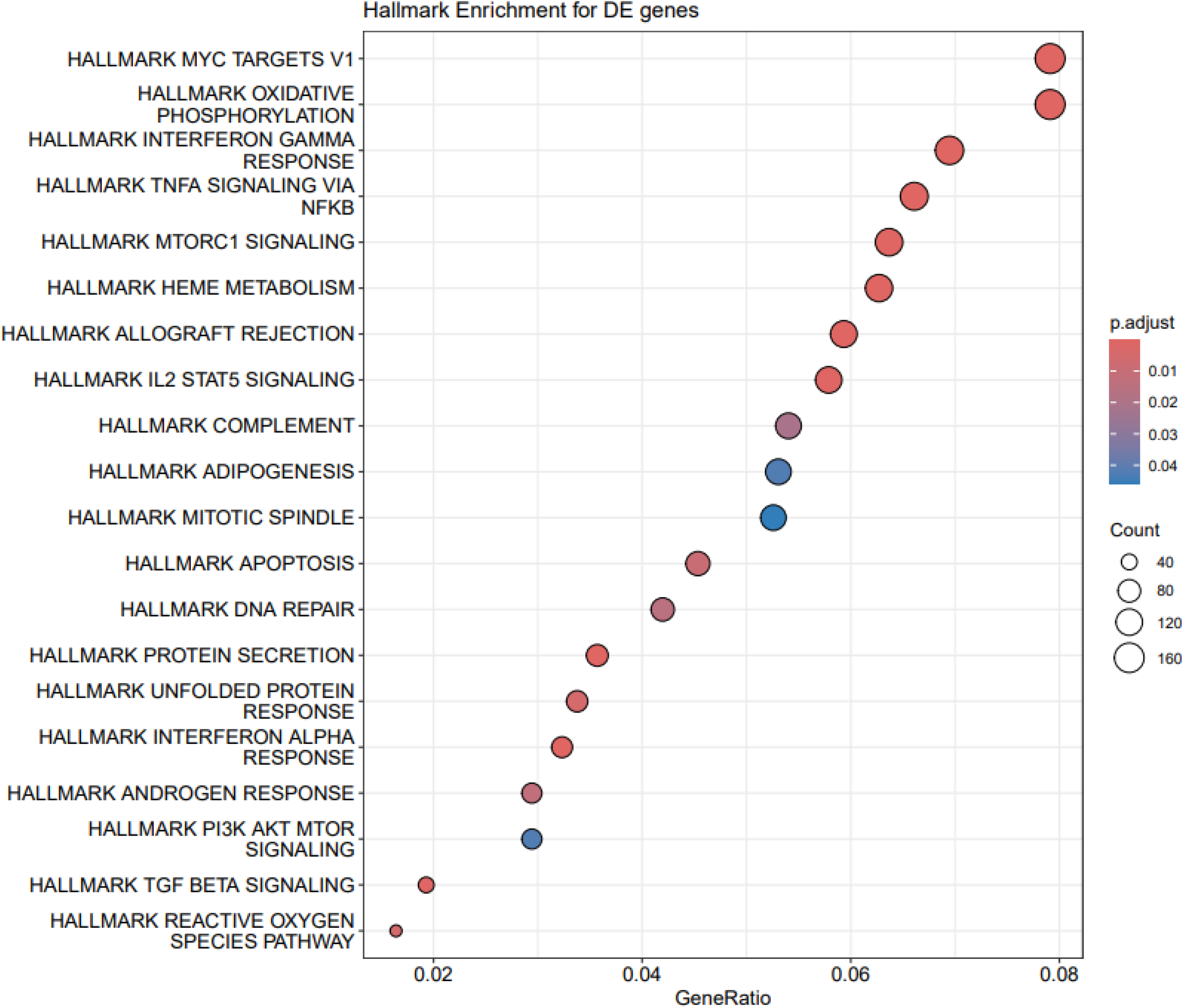
Hallmark Overrepresentation Analysis for DE genes

